# Exercise Reverses the Sedentary Cardiac Phenotype in Obesity: A Systematic Review and Meta-Analysis of Hemodynamic, Structural, and Functional Adaptations

**DOI:** 10.64898/2026.02.25.26347138

**Authors:** Ahmet Kurtoğlu, Musa Türkmen, Ertuğrul Kurtoğlu, Bekir Çar

## Abstract

Sedentary lifestyle and obesity are considered to be significant risk factors that create a pathway for the appearance of the ‘sedentary cardiac phenotype’ consisting of cardiac atrophy, myocardial stiffening, and altered haemodynamics. Although exercise training has the potential to reverse this detrimental process, the literature data on the magnitude of improvements and the certainty of evidence are inconsistent. This systematic review and meta-analysis aimed to evaluate the effects of exercise interventions on cardiac morphology, systolic/diastolic function, and haemodynamics in sedentary and obesity-prone individuals.

**Method:** In accordance with the PRISMA guidelines, the study was conducted by searching the PubMed, Web of Science, and Scopus databases from 1990 to 2025 without applying any filters, using Covidence software. As a result of this comprehensive search, 15 randomised controlled trials (RCTs; N=559) comparing exercise training with a control group in sedentary individuals were included in the analysis. Data were pooled using the Standardised Mean Difference (SMD) and a random-effects model. Publication bias and methodological robustness of the results were tested using the Egger regression test, the Trim-and-Fill method, and Leave-One-Out sensitivity analysis. The certainty of the evidence was graded using the GRADE system.

**Results:** Exercise training was associated with a significant reduction in resting HRs and SBPs, which was a strong improvement in the haemodynamic profile. The improvements in SV and LVEF, although on the statistical threshold in the primary analysis, were statistically significant and methodologically stable in the Leave-One-Out sensitivity analysis, which excluded confounding studies. The exercise training was associated with a marked improvement in the E/A ratio and S’ wave, and the triggering of a physiological athlete’s heart-like eccentric hypertrophy, defined by improvements in LVMass and LVEDV. The exercise training was associated with diastolic adaptation and mass increase, with HIIT being the most superior method for diastolic adaptation and mass increase, and aerobic exercise being the most effective method for blood pressure reduction. Importantly, the meta-regression analyses revealed two important findings: first, the improvement in blood pressure and diastolic function was independent of weight loss; second, the improvement in structure and function was linearly related to improvements in body composition.

**Conclusion:** Exercise acts as a ‘cardiac polypill’ reversing the sedentary phenotype by improving hemodynamics and diastolic function independently of weight loss, while linking structural remodeling to BMI optimization; our data prioritize HIIT for structural/diastolic gains and Aerobic training for blood pressure control.

## INTRODUCTION

Failure to meet minimum activity guidelines and a sedentary lifestyle, which means occasional, random participation in physical activities ^1^ has become more pronounced with the increase in factors that make our lives easier ^2^. A sedentary lifestyle can be said to be one of the greatest public health problems of our time ^3^. In particular, the weakening of physical health due to inactivity invites an increase in diseases such as obesity and type-2 diabetes, as well as various health problems such as coronary heart disease or hypertension, which result in disability or death ^3,4^.

Regular physical activity plays an important role in maintaining a healthy lifestyle and both preventing and improving various illnesses ^5–7^. It is known that the risk factors are significantly reduced, particularly due to cardiovascular adaptations resulting from long-term regular exercise ^8,9^. To understand these protective effects of exercise on cardiac morphology and haemodynamics, studies have focused primarily on the left ventricle (LV), with parameters such as left ventricular end-diastolic volume (LVEDV) being considered an important guide in determining low cardiorespiratory fitness ^10^. A review of the literature reveals numerous primary studies focusing on the effects of different exercise types on the LV in sedentary individuals. For example, step aerobic exercise has been reported to improve structural parameters such as left ventricular end-diastolic diameter (LVEDD), intraventricular septal thickness (IVS), and left ventricular posterior wall thickness (LVPW), as well as systolic and diastolic function parameters. Furthermore, while core exercise has been reported to improve LVEDD structural parameters along with systolic function ^11^,, structured endurance ^12^ and resistance exercises have been reported to cause an increase in LVEDV ^13^.

Additionally, high-intensity interval training (HIIT) has been found to improve the E/A ratio, an indicator of diastolic function, and stroke volume, while moderate-intensity continuous training (MICT) has been found to increase relative wall thickness (RWT) and aerobic power ^14^. However, the fact that these studies show significant differences in terms of protocol, duration and participant characteristics (gender, age, obesity) may lead to marked heterogeneity in the results. This situation makes it difficult to reach a clear conclusion regarding the relative effectiveness of different exercise modalities on the left ventricle and causes apparent inconsistencies between the findings. Meta-analysis studies conducted within the scope of this subject focus more on patients ^15,16^ and athlete ^17,18^ while a few studies have focused solely on endurance exercises ^19,20^.

In this context, the primary objective of the current systematic review and meta-analysis is to quantitatively synthesise the chronic effects of different exercise modalities (HIIT, Aerobic, Resistance and Team Sports) on cardiac morphology, diastolic function, and haemodynamic profile, and to reveal the physiological modification capacity of the “sedentary cardiac phenotype”. The primary hypothesis of our study is that cardiac remodelling is not a homogeneous process; structural hypertrophy (LVMass), filling dynamics (E/A) and autonomic balance (HR, Blood Pressure) will exhibit independent dose-response relationships depending on the type, intensity, weekly frequency and total duration of exercise. Furthermore, to test the frequently debated “necessity of weight loss” paradigm in obesity management, a secondary mechanistic hypothesis will be examined: whether cardiovascular adaptations progress independently of or in parallel with BMI changes. Supported by advanced sensitivity analyses and meta-regression models, this study aims not only to resolve the heterogeneity in the literature but also to provide a clinical basis for developing evidence-based and targeted exercise prescriptions that maximise cardiovascular protection in sedentary and obesity-prone populations.

## METHOD

### Registration of Systematic Review and Meta-Analysis Protocol

This systematic review and meta-analysis study was conducted in accordance with the Preferred Reporting Items for Systematic Reviews and Meta-Analyses (PRISMA) guidelines ^21,22^. The PRISMA checklist for this study is included as electronic supplementary material in Table S1. The study protocol has been registered in the International Prospective Register of Systematic Reviews (PROSPERO) system under the number CRD420261292046.

### Eligibility Criteria

Peer-reviewed journal articles written in English or Turkish that examine the effect of exercise interventions on left ventricular hypertrophy in sedentary individuals who occasionally engage in physical activity in a random manner that does not meet minimum activity guidelines have been considered ^1^. Review or descriptive articles, conference proceedings, and studies with samples consisting only of athletes or patients who have had myocardial infarction or high hypertension were excluded. In addition, no restrictions were applied in terms of country, time period, or study design. Studies that included left ventricular hypertrophy among cardiac parameters were included. The inclusion and exclusion criteria for this study are shown in Table 1.

**Table 1.**
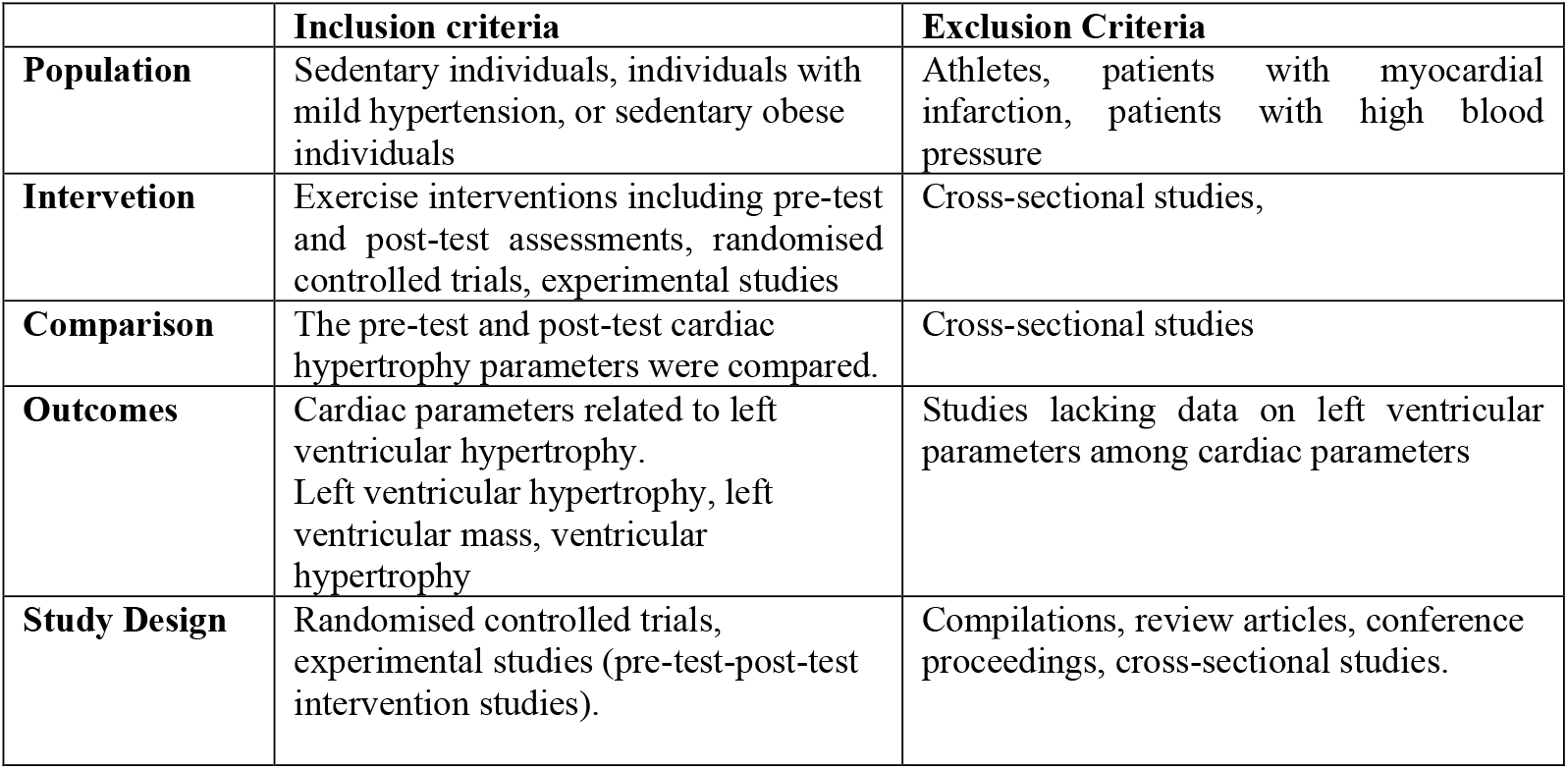
Inclusion and exclusion criteria according to the PICOS method.

|  | **Inclusion criteria** | **Exclusion Criteria** |
| --- | --- | --- |
| **Population** | Sedentary individuals, individuals with mild hypertension, or sedentary obese individuals | Athletes, patients with myocardial infarction, patients with high blood pressure |
| **Intervention** | Exercise interventions including pre-test and post-test assessments, randomised controlled trials, experimental studies | Cross-sectional studies, |
| **Comparison** | The pre-test and post-test cardiac hypertrophy parameters were compared. | Cross-sectional studies |
| **Outcomes** | Cardiac parameters related to left ventricular hypertrophy. Left ventricular hypertrophy, left ventricular mass, ventricular hypertrophy | Studies lacking data on left ventricular parameters among cardiac parameters |
| **Study Design** | Randomised controlled trials, experimental studies (pre-test-post-test intervention studies). | Compilations, review articles, conference proceedings, cross-sectional studies. |

### Search Strategy and Selection of Studies

To reach the final number of articles to be included in this study, a comprehensive search was conducted using the following keywords without applying any other filters from seven different search engines—PubMed, Web of Science and Scopus —from 1990 to the present date of 2025. These searches were performed using the Covidence program.

The search syntax was created by combining free-text terms found within the controlled vocabulary (MeSH - Medical Subject Headings) and the title/abstract ([tiab] or TS=Topic). Specifically, the search string consists of: (1) Population (‘sedentary’, ‘physical inactivity’, ‘untrained’), (2) Intervention (‘aerobic training’, ‘resistance training’, ‘HIIT’, ‘exercise training’), (3) Outcome Variables (‘echocardiography’, ‘cardiac remodeling’, ‘left ventricular mass’ and derivatives), and (4) Study Design (‘cohort’, ‘longitudinal’, ‘prospective’) keywords integrated using Boolean operators (AND/OR).

Details regarding the articles included in the study are provided in Figure 1. The PRISMA flow diagram was obtained using Covidence software. This software has a very practical application for eliminating duplicate studies and is a time-efficient tool. During this process, the articles obtained by two independent authors (AK and MT) were first reviewed by their titles and abstracts, and disagreements were resolved through consensus. Subsequently, the full texts of the articles were reviewed, and 15 articles meeting the exclusion and inclusion criteria were included in the study. The full texts of these studies were uploaded to the Covidence software.

**Figure 1.**
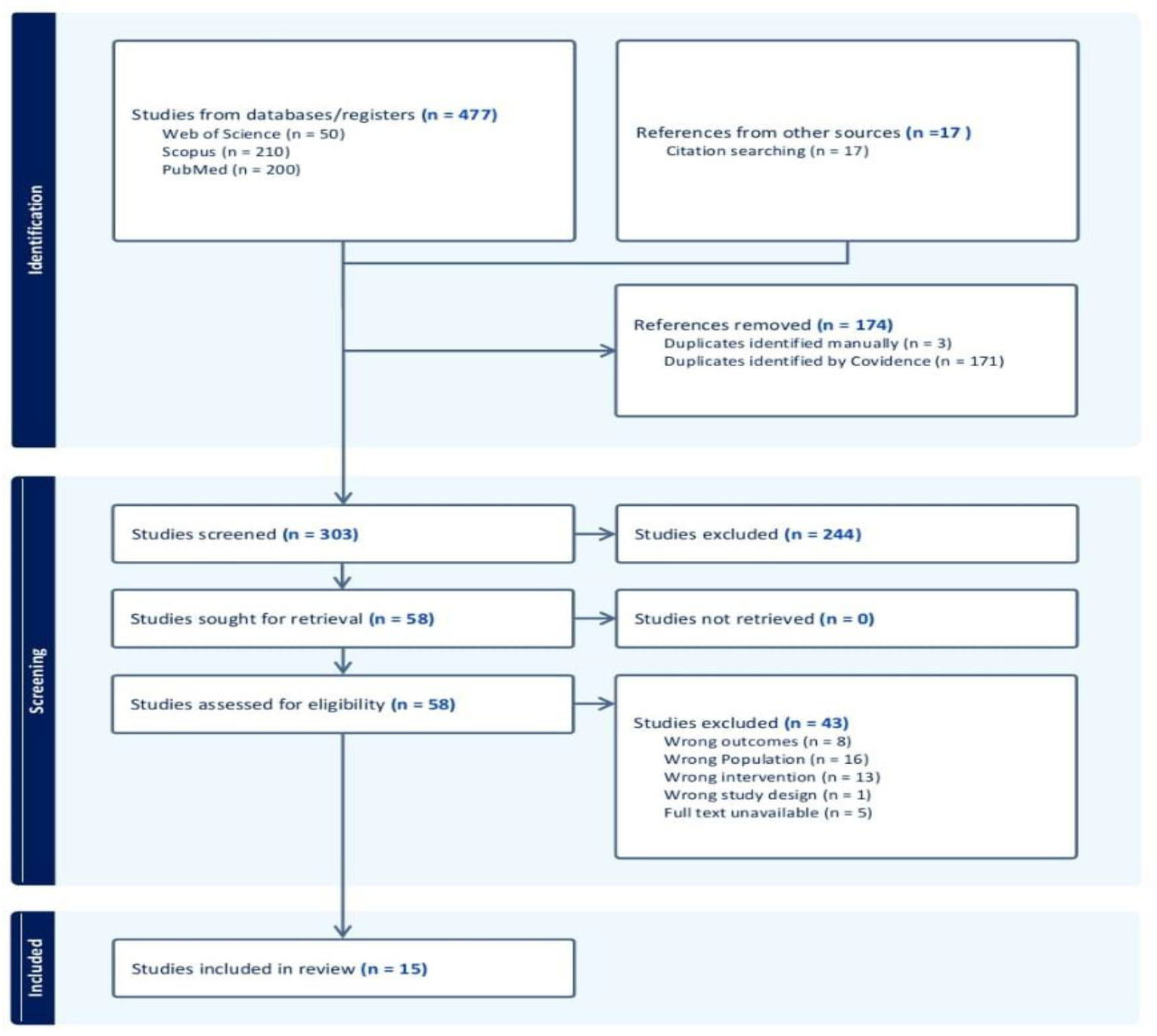
PRISMA flow diagram.

### Data Extraction Process

After the final studies were determined, the data extraction phase was initiated. The data extraction form was prepared based on the Data Extraction and Assessment Form in the Cochrane Handbook ^23^. After both researchers collected the data, disagreements were resolved through compromise and the data were combined. During the data extraction process of the study, the means and standard deviations related to the dependent variables were transferred to a Microsoft Excel spreadsheet, and the following headings were determined for the other data: Study year, country where the study was conducted, study design, number of participants, participant characteristics, exercise type, exercise intensity, weekly exercise frequency, single session duration, total duration, weekly exercise volume, average age of participants, BMI values, smoking rate among participants, diabetes mellitus rate, SBP, DBP, HR, LVEDV, LVESV, LVEDD, LVESD, EF, IVSd, IVSs, PWT, LVM, LAD, LAV, E, A, S, E’, A’, S’, E/A, E/E’, IVRT, IVCT, and SV average values.

Some data from the studies obtained are presented graphically. Since the findings obtained in these studies do not have numerical values, the Web Plot Digitizer software tool was used to overcome this problem and obtain numerical data ^24^ (https://automeris.io/ erişim tarihi: 17.01.2026).

### Quality Assessment

The Risk of Bias 2 (RoB 2) tool developed by The Cochrane Collaboration was used to assess the methodological quality and risk of bias of the articles included in the study. Table 2 presents the RoB 2 assessment results for the studies included in the research. This tool evaluates articles in five main areas from a methodological perspective (bias arising from randomization, bias arising from planned interventions, bias arising from incomplete outcome data, bias arising from outcome measurement, bias arising from reporting of outcomes). The results obtained from the evaluation of these areas help us understand whether the results of the studies can be trusted ^25^.

**Table 2.** The RoB 2 assessment results of the studies.

| Authors and Date | 1 | 2 | 3 | 4 | 5 | Overall |
| --- | --- | --- | --- | --- | --- | --- |
| Andersen L.J. et al.,2010 | Low risk | Some concerns | Low risk | Low risk | Low risk | Some concerns |
| Andersen L.J. et al.,2014 | Low risk | Some concerns | Low risk | Low risk | Low risk | Some concerns |
| Baglivo et al., 1990 | High risk | High risk | Low risk | Some concerns | Some concerns | High risk |
| Howden et al., 2018 | Low risk | Low risk | Low risk | Low risk | Low risk | Low risk |
| Huang et al., 2019 | Low risk | Low risk | Low risk | Low risk | Low risk | Low risk |
| Lekavich et al.,2021 | Low risk | Low risk | Low risk | Low risk | Low risk | Low risk |
| McKenna et al., 2023 | Low risk | Low risk | Low risk | Low risk | Low risk | Low risk |
| Millen et al., 2014 | Low risk | Low risk | Low risk | Low risk | Low risk | Low risk |
| Norha et al., 2025 | Low risk | Some concerns | Low risk | Low risk | Low risk | Low risk |
| Oxborough et al., 2019 | Low risk | Low risk | Low risk | Low risk | Low risk | Low risk |
| Scharf et al.,2015 | Low risk | Low risk | Some concerns | Low risk | Low risk | Some concerns |
| Schmidt et al., 2014 | Low risk | Low risk | Low risk | Low risk | Low risk | Low risk |
| Sjúrðarson et al., 2024 | Low risk | Low risk | Low risk | Low risk | Low risk | Low risk |
| Spence et al., 2011 | Low risk | Low risk | Low risk | Low risk | Low risk | Low risk |
| Zheng et al., 2018 | Low risk | Low risk | Some Concerns | Low risk | Low risk | Some concerns |
| Domain 1: Randomisation process, Domain 2: Alterations from planned interventions, Domain 3: Missing outcome data, Domain 4: Measurement of outcome, Domain 5: Selection of the reported result |  |  |  |  |  |  |

**Table 3.** The characteristics of the included studies.

| Study ID | Year | Country | Exercise Type | n | Age (Years) | Male (%) | BMI (kg/m<sup>2</sup>) | Diabetes (%) | Smoking (%) | Intervention Protocol | Intensity |
| --- | --- | --- | --- | --- | --- | --- | --- | --- | --- | --- | --- |
| <sup>35</sup> | 2010 | Denmark | Running | 18 | 36.5 ± 8.2 | %0 | 24.3 ± 4.1 | NaN | NaN | 16 weeks; 1.9 days/wk; 60 min/session | Moderate |
|  |  |  | Football | 19 |  |  |  |  |  | 16 weeks; 1.8 days/wk; 60 min/session | High |
|  |  |  | Control | 10 |  |  |  |  |  | Standard Care / No Exercise | - |
| <sup>36</sup> | 2021 | USA | Resistance | 10 | 47 ± 12.3 | %30 | 29.3 ± 2.3 | %0 | - | 24 weeks; 3 days/wk; NA min/session | Moderate+High |
|  |  |  | Aerobic | 11 | 53.1 ± 6.4 | %64.3 | 31.7 ± 3.4 | %0 | - | 24 weeks; NA days/wk; NA min/session | Moderate+High |
| <sup>37</sup> | 2023 | USA | Control | 10 | 34 ± 9 | %50 | 28.2 | - | - | Standard Care / No Exercise | - |
|  |  |  | Aerobic+ Resistance | 19 | 43 ± 13 | %57.9 | 28.2 | - | - | 24 weeks; NA days/wk; NA min/session | - |
| <sup>38</sup><br>A12 | 2014 | South Africa | Aerobic | 16 | 44.2 ± 5.7 | %69 | 31.8 ± 3.3 | - | %0 | 6 weeks; 3 days/wk; 83 min/session | Moderate+High |
|  |  |  | Aerobic | 16 | 44.9 ± 4.5 | %75 | 30.6 ± 5.1 | - | %0 | 6 weeks; 3 days/wk; 83 min/session | Moderate+High |
| <sup>39</sup> | 2025 | Finland | Control | 31 | 52.7 ± 7.5 | %100 | 31.7 ± 4.6 | - | %0 | Standard Care / No Exercise | - |
|  |  |  | Aerobic | 33 | 59.3 ± 6 | %100 | 31.5 ± 4 | - | %0 | NA weeks; NA days/wk; NA min/session | Low+moderate |
| <sup>40</sup> | 2019 | UK<br>Australia<br>Holland | Resistance | 13 | 26.6 ± 1.3 | %100 | 24.7 ± 1 | - | %0 | 24 weeks; 3 days/wk; 60 min/session | High |
|  |  |  | Endurance | 10 | 28.4 ± 1.9 | %100 | 24.2 ± 1.3 | - | %0 | 24 weeks; 3 days/wk; 60 min/session | High |
| <sup>41</sup> | 2015 | Germany | HIIT | 42 | 44.2 ± 4.8 | %100 | 28.1 ± 3.5 | %5 | %10 | 16 weeks; NA days/wk; NA min/session | High |
|  |  |  | Control | 42 | 42.4 ± 5.5 | %100 | 27 ± 4.1 | %5 | %10 | Standard Care / No Exercise | - |
| <sup>42</sup> | 2014 | Denmark | Football | 21 | 45.8 ± 7.2 | %100 | 30.3 ± 3.3 | - | %20 | 24 weeks; 2 days/wk; 60 min/session | High |
|  |  |  | Control | 10 | 46.9 ± 7.6 | %100 | 29.6 ± 3.6 | - | %18.2 | Standard Care / No Exercise | - |
| <sup>43</sup> | 2014 | Denmark | Resistance | 9 | 69.1 ± 3.1 | %100 | 27.4 ± 2.8 | %0 | %0 | 52 weeks; 3 days/wk; 60 min/session | Moderate+High |
|  |  |  | Football | 9 | 68 ± 4 | %100 | 26.1 ± 3.9 | %0 | %0 | 52 weeks; 3 days/wk; 60 min/session | High |
|  |  |  | Control | 8 | 67.4 ± 2.7 | %100 | 27.9 ± 4.6 | %0 | %0 | Standard Care / No Exercise | - |
| <sup>44</sup> | 2024 | Denmark | Swimming_moderate | 21 | 46.9 ± 3 | %0 | 30.7 ± 5.7 | - | - | 15 weeks; 3 days/wk; 60 min/session | Moderate |
|  |  |  | Swimming_high | 21 | 44.1 ± 4.7 | %0 | 28.8 ± 3.7 | - | - | 15 weeks; 3 days/wk; 25 min/session | High |
|  |  |  | Control | 20 | 43.7 ± 4.3 | %0 | 28 ± 4.1 | - | - | Standard Care / No Exercise | - |
| 45 | 2011 | Australia | Resistance | 13 | 26.6 ± 1.3 | %100 | 24.7 ± 1 | - | - | NA weeks; 3 days/wk; 60 min/session | Moderate+High |
|  |  |  | Endurance | 10 | 28.4 ± 1.9 | %100 | 24.2 ± 1.3 | - | - | 24 weeks; 3 days/wk; 60 min/session | Low+Moderate+High |
| 46 | 2019 | China | Tai Chi | 85 | 61 ± 5.2 | %32.9 | - | %9.4 | - | 12 weeks; 5 days/wk; 60 min/session | Mild+moderate |
|  |  |  | Control | 85 | 60.7 ± 6 | %28.2 | - | %12.9 | - | Standard Care / No Exercise | - |
| 47 | 1990 | Argentina | Control | 15 | 49.3 ± 3.2 | %66.7 | - | - | - | Standard Care / No Exercise | - |
|  |  |  | Aerobic | 17 | 50.8 ± 7.9 | %70.6 | - | - | - | 68 weeks; 3 days/wk; 50 min/session | Moderate |
| 48 | 2018 | USA | Control | 28 | 51.4 | %46 | 26.2 | - | %0 | Standard Care / No Exercise | - |
|  |  |  | Aerobik+Resistance | 33 | 53.2 | %45 | 25.8 | - | %0 | 104 weeks; 4 days/wk; 60 min/session | Low+moderate+high |
| 49 | 2019 | Taiwan | MIIT | 18 | 21.9 ± 0.6 | %100 | 22.6 ± 0.7 | - | %0 | 6 weeks; 5 days/wk; 30 min/session | Moderate |
|  |  |  | HIIT | 18 | 21.4 ± 0.4 | %100 | 22.7 ± 0.6 | - | %0 | 6 weeks; 5 days/wk; 30 min/session | High |
|  |  |  | Control | 18 | 22 ± 0.5 | %100 | 21.8 ± 0.8 | - | %0 | Standard Care / No Exercise | - |

### Statistical Analyses

All statistical analyses and data visualisations were performed using the R Statistical Software Environment (R Foundation for Statistical Computing, Vienna, Austria). The ‘meta’ ^26^ and “metafor” ^27^ packages were used for meta-analysis procedures, while the ‘dmetar’ library was used for publication bias and sensitivity analyses. The analysis process was conducted in full compliance with the guidelines of the Cochrane Handbook for Systematic Reviews of Interventions ^23^.

The mean change values and standard deviations (SD) before and after the intervention were used for continuous variables (e.g., LVEF, LVMass, E/A ratio, BMI). To normalise measurement methods and scale differences between studies, the Standardised Mean Difference (SMD) and 95% Confidence Interval (CI) were calculated as effect size metrics ^28^. Due to the expected clinical and methodological heterogeneity between exercise protocols and participant populations (sedentary, at risk of obesity, etc.), the Random-Effects Model and Inverse Variance method, which offer a more conservative approach than fixed effects, were preferred in all pooling procedures ^29^.

To investigate inter-study statistical heterogeneity, the Cochran Q test (p < 0.10) and the I^2^ statistic, which measures the percentage of variation due to factors other than chance, were used. The level of heterogeneity between studies was defined as low, moderate, and high for an I^2^ statistic value between 33%, between 33% and 66%, and above 66%, respectively ^30^. To explore possible reasons for heterogeneity (e.g., influence independent of BMI, dose-response relationship), subgroup analysis for categorical variables (Exercise Type: HIIT/Aerobic) and univariate meta-regression analysis for continuous variables (Total Weeks, Frequency, BMI Change) were carried out.

To test the methodological robustness of the findings and determine whether the overall result of a single study was artificially influenced (influential cases), a “Leave-One-Out” sensitivity analysis was applied. In this procedure, one study was removed from the pool at each step, and the overall effect size and statistical significance (p-value) were iteratively recalculated ^27^.

Publication bias risk was examined using a multidimensional approach. Visual asymmetry was assessed using Funnel Plots; statistical asymmetry was tested using Egger’s linear regression test and Begg’s rank correlation test (p < 0.05 was considered significant) ^31^. Rosenthal’s Fail-Safe N (FSN) number was calculated to measure the robustness of the results against unpublished “negative” studies ^32^. For parameters with asymmetry detected in the funnel plot (e.g., HR, E/A), Duval and Tweedie’s “Trim-and-Fill” method was applied to estimate the adjusted effect size by simulating missing studies ^33^.

The certainty level of evidence (Certainty of Evidence) obtained for each outcome was assessed using the GRADE approach (Grading of Recommendations Assessment, Development and Evaluation) and rated as High, Moderate, Low, or Very Low based on the risk of error, inconsistency (I^2^), indirectness, uncertainty, and publication bias ^34^. In all analyses, the statistical significance level was set at p < 0.05 (two-tailed).

## RESULTS

This meta-analysis included a total of 15 studies conducted between 1990 and 2025 ^47^ and ^39^). The geographical distribution of the studies is quite widespread and includes data from European countries such as Denmark, Finland, Germany, the UK, and the Netherlands; North American countries such as the USA; Asian countries such as China and Taiwan; South American countries such as Argentina; and the African country of South Africa.

From the demographic data of the cohorts included in the studies, it is clear that the studies included participants from a broad spectrum of ages, from young adults (∼21 years; ^49^) to geriatric patients (∼69 years; ^43^). The gender distribution of the participants in the included studies is quite variable. Studies (Andersen et al., 2014; Huang et al., 2019; Norha et al., 2025; Oxborough et al., 2019; Scharf et al., 2015) and ^43^ included only males, while the swimming intervention study conducted exclusively with female participants is denoted by ^44^. The body mass index (BMI) of the participants included in the studies is mostly in the overweight or obese category [^36,38,39^; BMI > 30 kg/m^2^], while the participants in the studies denoted by ^49^ and ^40^ are of normal weight. As regards comorbid conditions, the participants with diabetes (^41,46^ and smokers ^41,42^ are included in certain studies.

Exercise protocols: The types of exercises included in the intervention protocols are quite heterogeneous. They can be divided into the following types: Aerobic Exercises: Running ^35^, cycling ^47^, swimming ^44^, and aerobic exercises in general ^36,38,39^.

Team Sports: Football training (Andersen et al., 2010, 2014; Schmidt et al., 2014). Resistance Exercises: Isolated or combined exercises (Lekavich et al., 2021; Oxborough et al., 2019; Schmidt et al., 2014; Spence et al., 2011).

High-Intensity Interval Training: ^41,49^.

Mind-Body Exercises: Tai Chi ^46^.

Duration of the exercises: The exercise protocols lasted from 6 weeks ^38,49^ to 104 weeks ^48^. The frequency of the exercises is from 1.8 days/week ^35^ to 5 days/week ^46,49^. The intensity of the exercises ranges from light-moderate intensity in the Tai Chi study ^46^ to high intensity in the studies denoted by the codes (Andersen et al., 2010, 2014; Oxborough et al., 2019; Scharf et al., 2015) and ^44^. The control participants in all the studies continued with standard care without any exercise intervention.

The current meta-analysis includes a total of 15 studies investigating the impact of the exercise protocols applied to sedentary individuals. The data obtained from the studies included in the current analysis comprise a total of 559 participants (Intervention and Control). The changes in the cardiac parameters were calculated using the Standardised Mean Difference (SMD) and the net % Change using the change from baseline scores.

### Haemodynamic Responses

Significant and clinically relevant improvements in the haemodynamic profile were achieved with exercise training. The exercise group recorded a highly significant 7.4% reduction in the resting HR compared with the control group (n=280; SMD = -1.09, p<0.001). Likewise, the exercise group also recorded a net decrease of 2.7% in systolic blood pressure (SBP) (SMD = -0.78, p = 0.015). In the analysis using the expanded dataset (n=310), stroke volume (SV), a major indicator of the pumping ability of the heart, was increased by 7.3% in the exercise group, recording a strong effect size (SMD = 1.43), but statistical significance was borderline due to increased heterogeneity (p = 0.053).

### Systolic Function

Left ventricular systolic function was studied multidimensionally. In the study where classical LVEF was used (n=559), a 1.0% positive result was found for the exercise group, although it failed to be statistically significant (p=0.053). In the study where the S’ wave, a more sensitive measure of contractility using TDI (n=122), was used, a remarkable result of a 13.9% increase in the exercise group was found (SMD=0.58; p=0.003). In addition to this, a significant increase in IVSs was found to be 3.5% (p=0.008). These results demonstrate the significant effect of exercise on the mechanics of the myocardium despite the stability of the systolic function.

### Diastolic Function

Significant improvements in diastolic parameters were recorded with exercise. IVRT (n=88) shortened by 18.9% in the exercise group (SMD = -1.06, p < 0.001), indicating a marked acceleration in left ventricular relaxation kinetics. The E/A ratio (n=446), reflecting left ventricular filling dynamics, showed a net increase of 14.0% in the exercise group (SMD = 0.76, p < 0.001), and the E wave amplitude increased by 4.0% (p = 0.033), supporting improved diastolic compliance.

### Structural Remodelling

Exercise training led to changes in left ventricular morphology consistent with the physiological ‘athlete’s heart’ adaptation. LVEDV (n=360) showed a statistically significant increase of 10.1% in the exercise group (SMD = 0.53, p = 0.004). Parallel to this, a statistically significant 6.7% increase in LVMas (n=396) was observed (SMD = 0.90, p = 0.022). The absence of significant differences between groups in terms of IVSd, PWT, and LVEDD suggests that the developing hypertrophy is physiological (eccentric) rather than pathological (concentric) in nature.

In the main analysis presented in Figure 2, despite the strong clinical effect size observed, particularly for SV and LVEF, statistical significance was borderline (p = 0.053). To further investigate these results, a Leave-One-Out sensitivity analysis was applied, which aims to test the methodological robustness of these results (Table 4). The results of the analysis indicated that, although there is a positive effect of exercise on LVEF and SV, there is heterogeneity due to specific studies. It was found that these results were being “masked” by specific studies, which were causing heterogeneity. It was found that, although LVEF did not reach statistical significance, it did reach statistical significance (p < 0.05) once studies that decreased the effect (e.g., A1, A11, A19) were removed one by one. Similarly, for SV, it was found that once a specific outlier study (A11) was removed, it did reach a level of statistical significance (p < 0.05), statistically confirming clinical inference that LVEF and SV increase or remain unchanged due to exercise. It was also found that, although there is a significant increase in LVMass and SBP, these results were dependent upon the inclusion of two highly influential studies (A9, A20), as once these studies were removed, there was no longer any significant effect. However, results for parameters such as HR, LVEDV, and E/A ratio were found to be robust, as they were not affected by the removal of any studies.

**Table 4.** Summary of the Leave-One-Out sensitivity analysis evaluating the effect of individual studies on pooled effect sizes and the robustness of findings.

| Parameter | k_Studies | Original P | Robustness_Status | Influential_Studies |
| --- | --- | --- | --- | --- |
| SBP | 4 | 0.015 | SENSITIVE (Significance Lost) | A4, A9 |
| HR | 6 | 0.000 | Robust / Stable | None |
| LVEDV | 9 | 0.004 | Robust / Stable | None |
| LVEDD | 11 | 0.146 | Robust / Stable | None |
| LVESD | 7 | 0.244 | Robust / Stable | None |
| EF | 12 | 0.051 | SENSITIVE (Significance Gained) | A1, A1, A2, A11, A19, A21 |
| IVSd | 8 | 0.146 | Robust / Stable | None |
| PWT | 7 | 0.333 | SENSITIVE (Significance Gained) | A23 |
| LVMass | 10 | 0.022 | SENSITIVE (Significance Lost) | A9, A20 |
| LAD | 4 | 0.472 | Robust / Stable | None |
| E | 7 | 0.033 | SENSITIVE (Significance Lost) | A1, A9 |
| A | 6 | 0.010 | SENSITIVE (Significance Lost) | A1 |
| E' | 5 | 0.138 | SENSITIVE (Significance Gained) | A20 |
| A' | 5 | 0.646 | Robust / Stable | None |
| S' | 5 | 0.002 | Robust / Stable | None |
| E/A | 10 | 0.000 | Robust / Stable | None |
| E/E' | 5 | 0.664 | Robust / Stable | None |
| IVRT | 3 | 0.000 | Robust / Stable | None |
| SV | 6 | 0.053 | SENSITIVE (Significance Gained) | A11 |

**Figure 2.**
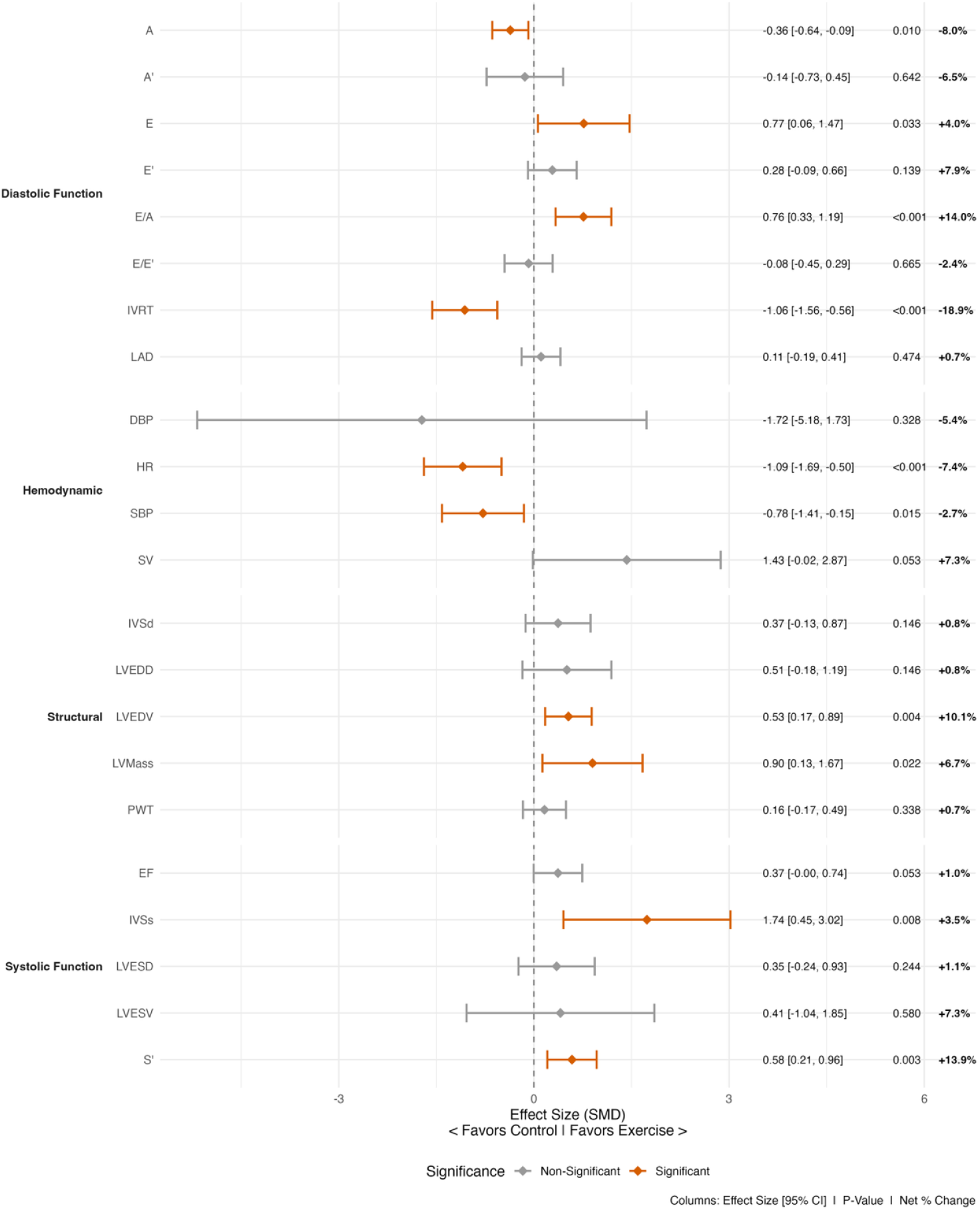
Summary forest plot showing the pooled effects of exercise interventions on cardiac structure and function parameters.

**Figure 2.**
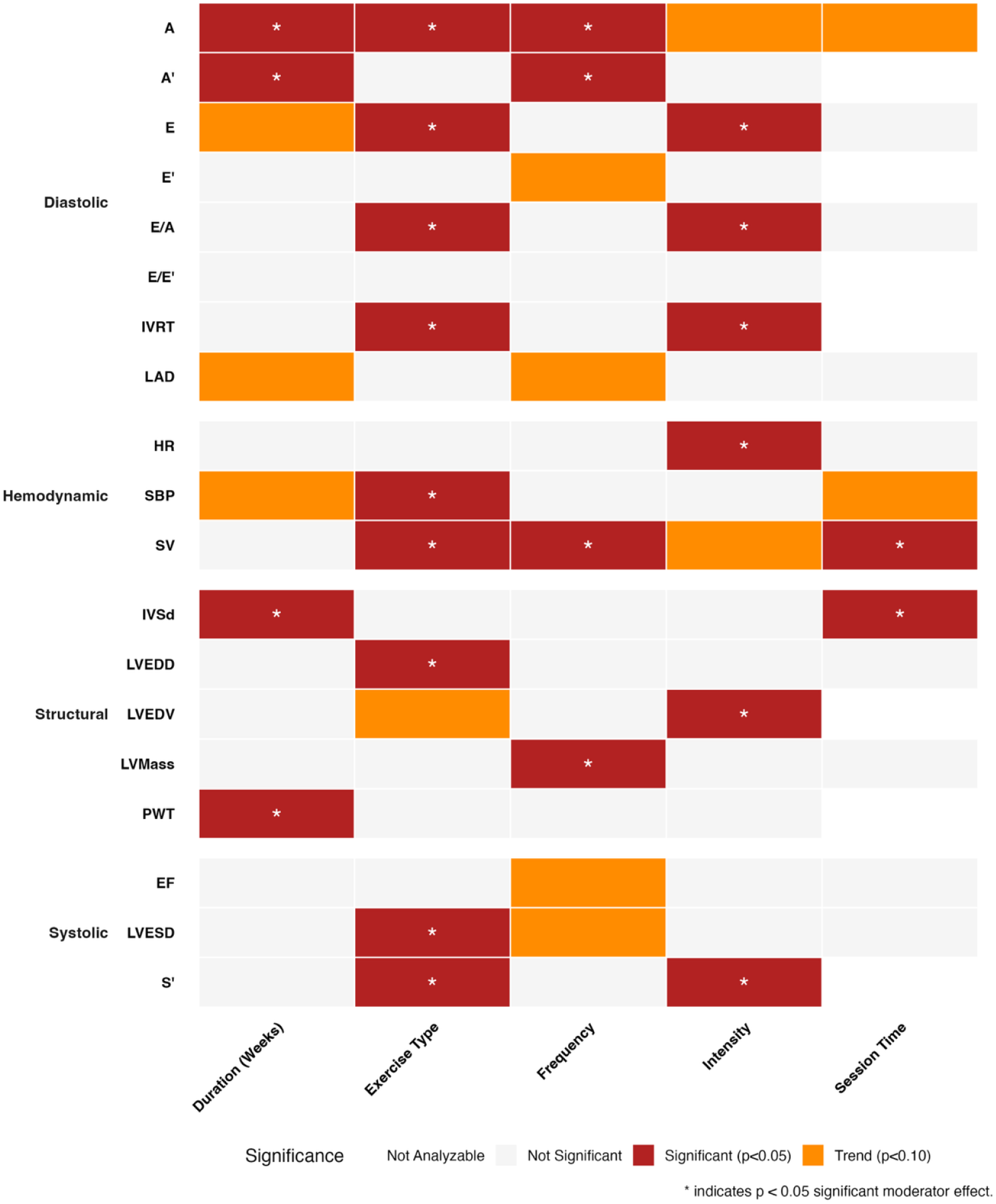
Moderator analysis matrix showing the effects of exercise prescription components on cardiac structure and function parameters.

Meta-regression and subgroup analysis performed to assess the decisive influence of the essential elements of the exercise programme, such as type, intensity, duration, and frequency, on cardiac parameters showed that structural and functional changes are influenced by different dynamics of exercise.

In relation to the analysis of the haemodynamic parameters, exercise modality was found to be an important cause of variation for SBP change (p = 0.002). Subgroup analysis showed that aerobic exercise protocols had the largest effect size (SMD = -0.40) for lowering blood pressure.

Change in resting HR was found to be influenced more by training intensity than training type (p < 0.001); high-intensity training was found to be statistically significant over moderate training for increasing parasympathetic tone. Team Sports was found to be an effective alternative for lowering HR (SMD= -0.54).

A dose-response relationship was found for S’ wave, an indicator of longitudinal systolic ventricular performance (p = 0.008); aerobic and high-intensity training were found to maximise myocardial contractility velocity (SMD = 0.75).

Although no statistical significance was found for the analysis of EF, an indicator of global systolic ventricular performance (p = 0.60), when examining the effect sizes, Resistance training was found to be the training method that increased EF to the largest extent (SMD = 1.06).

Similarly, for LVESD, an indicator of systolic wall tension, HIIT was found to be the training method that provided the strongest effect (SMD = 2.21).

One of the most interesting results of the study is the sensitivity of diastolic parameters to the type of exercise. A very strong moderating effect of the modality of the exercise on the E/A ratio, which reflects the dynamics of the filling of the left ventricle, was detected. HIIT protocols dramatically surpass the effects of Aerobic and Resistance exercises on diastolic compliance. In addition, HIIT protocols also demonstrated a dominant superiority compared to the other types of exercises regarding the increase of the early diastolic flow (E wave). On the contrary, regarding the IVRT, which reflects the active relaxation of the heart, aerobic exercises were identified as the most effective method to increase the speed of the relaxation kinetics. This implies the existence of specific responses of the phases of diastolic heart function to the various types of exercises. Positive effects of Team Sports (SMD > 0.45) were detected on the LAD and E’ parameters. The physiological increase of the LVMass was observed to have a positive and significant linear relationship with the frequency of the exercises performed during the week. In addition, the method of HIIT was the one which triggered the greatest increase of the mass. The most significant effect on the LVEDV and LVEDD was observed to be achieved through the application of the HIIT method and the implementation of high-intensity exercises. A significant effect of the total intervention duration (Total Weeks) on the structural wall thicknesses IVSd and PWT was detected. More specifically, the effect of the total intervention duration on the IVSd and PWT was observed to be decreasing over time.

Thus, in conclusion, it can be said that the analysis has revealed that, while the increase in functional capacity, i.e., diastolic E/A and systolic S’, is largely modulated by the intensity and type of exercise, particularly HIIT, the structural ‘athlete’s heart’ adaptations, i.e., LVMass and PWT, have been found to have a statistically stronger association with the frequency and duration of training.

The results of the meta-regression analysis, which was carried out to assess whether the cardiac improvements observed after exercise were dependent on the initial BMI of the participants or the change in BMI during the process, showed an evident difference between the functional and structural adaptations (Table 5).

**Table 5.** BMI and Cardiac Adaptation Relationship: Meta-Regression Analysis.

| Parameter | Studies | Beta_BMI | P_Value | Relationship |
| --- | --- | --- | --- | --- |
| SBP | 3 | 0,333891873 | 0,183 | Non-Significant |
| HR | 5 | 0,168056262 | 0,001 | Significant |
| LVEDV | 8 | -0,102872179 | 0,238 | Non-Significant |
| LVEDD | 9 | -0,252527527 | 0,020 | Significant |
| LVESD | 6 | -0,092556377 | 0,362 | Non-Significant |
| EF | 10 | -0,145604762 | 0,037 | Significant |
| IVSd | 7 | -0,162227991 | 0,040 | Significant |
| PWT | 5 | -0,030985764 | 0,690 | Non-Significant |
| LVMass | 10 | -0,330872172 | 0.000 | Significant |
| LAD | 3 | 0,14342324 | 0,522 | Non-Significant |
| E | 5 | -0,212976102 | 0,051 | Non-Significant |
| A | 5 | -0,046178151 | 0,292 | Non-Significant |
| E' | 4 | 0,032181521 | 0,817 | Non-Significant |
| A' | 4 | 0,04889499 | 0,788 | Non-Significant |
| S' | 4 | -0,092513491 | 0,312 | Non-Significant |
| E/A | 8 | -0,108991848 | 0,129 | Non-Significant |
| E/E' | 4 | 0,012026928 | 0,892 | Non-Significant |
| SV | 6 | -0,39633748 | 0,020 | Significant |

The most interesting outcome of the analysis results is that the reduction in SBP did not reveal any statistically significant relationship with the BMI parameters, i.e., (β = 0.33, p = 0.183). This demonstrates that the antihypertensive effect of exercise training is independent of the loss of body weight or the level of obesity; thus, exercise regulates blood pressure even in sedentary individuals without any alteration in body weight. Moreover, the diastolic parameters, i.e., E/A, E’, A’, IVRT, which reflect the left ventricular filling pressures and the relaxation properties, were not found to be altered by the BMI modulations (p > 0.05).

In contrast to functional parameters, the results of the analysis of cardiac structural adaptations demonstrated a significant interaction of the effect of exercise training on left ventricular mass (LVMass, p < 0.001), left ventricular diameter (LVEDD, p = 0.020), and stroke volume (SV, p = 0.020) with BMI. The negative value of the regression coefficients of the effect of exercise training on left ventricular mass (LVMass, β = -0.33) and stroke volume (SV, β = -0.39) indicates that high BMI values or low weight reduction modulate the effect of exercise training. In addition, the analysis of the results demonstrated that the effect of exercise training on systolic/autonomic parameters such as heart rate (HR, p = 0.001) and ejection fraction (EF, p = 0.037) was parallel to the change in BMI.

Thus, the results of the analysis of the effect of exercise training on cardiovascular parameters demonstrated that the effect of exercise training is associated with a dual mechanism of action: the reduction of blood pressure and the maintenance of diastolic function in obese sedentary subjects, irrespective of weight reduction (Cardiac Risk Reduction). In addition, the results of the analysis of the effect of exercise training on cardiovascular parameters demonstrated that the effect of exercise training is associated with the strengthening of the heart, as indicated by the increase in left ventricular mass (LVMass) and stroke volume (SV); however, these parameters are related to the improvement of body composition (BMI optimisation).

The risk of publication bias for the studies included in the analysis was also evaluated in terms of the robustness of the results using Egger’s regression test, Begg’s rank correlation method, and Rosenthal’s fail-safe N (FSN) tests in Table 6. The results of the analysis showed that the parameters of interest such as SV, LVMass, and E/A ratio had relatively high values of FSN equal to 165, 151, and 135, respectively. This clearly proves that the results of the observed effect of exercise on cardiac performance are not random and are resistant to the hypothetical influence of negative studies that have not been published. Although the Egger’s test and Begg’s rank correlation tests for HR, E wave, and E/A ratio showed the presence of a potential asymmetry in the funnel plot for these parameters (p < 0.05), the results of the sensitivity analysis using the Trim-and-Fill method, which was performed to confirm that the observed asymmetry was not related to publication bias, showed that no studies were assigned for the above-mentioned parameters. This clearly proves that the observed asymmetry is related to the variability of results or the small-study effect rather than the absence of studies. Moreover, it was also observed that the limited corrections for the results of the analysis for parameters such as LVEDV and LAD did not influence the results of the effect size. As a result of this analysis, it was clearly demonstrated that the results of the present meta-analysis have a high level of representativeness of the literature data and do not reflect the presence of significant publication bias.

**Table 6.**
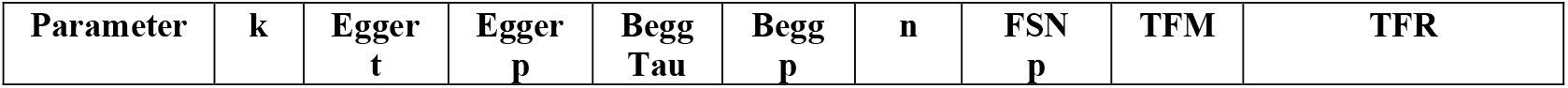
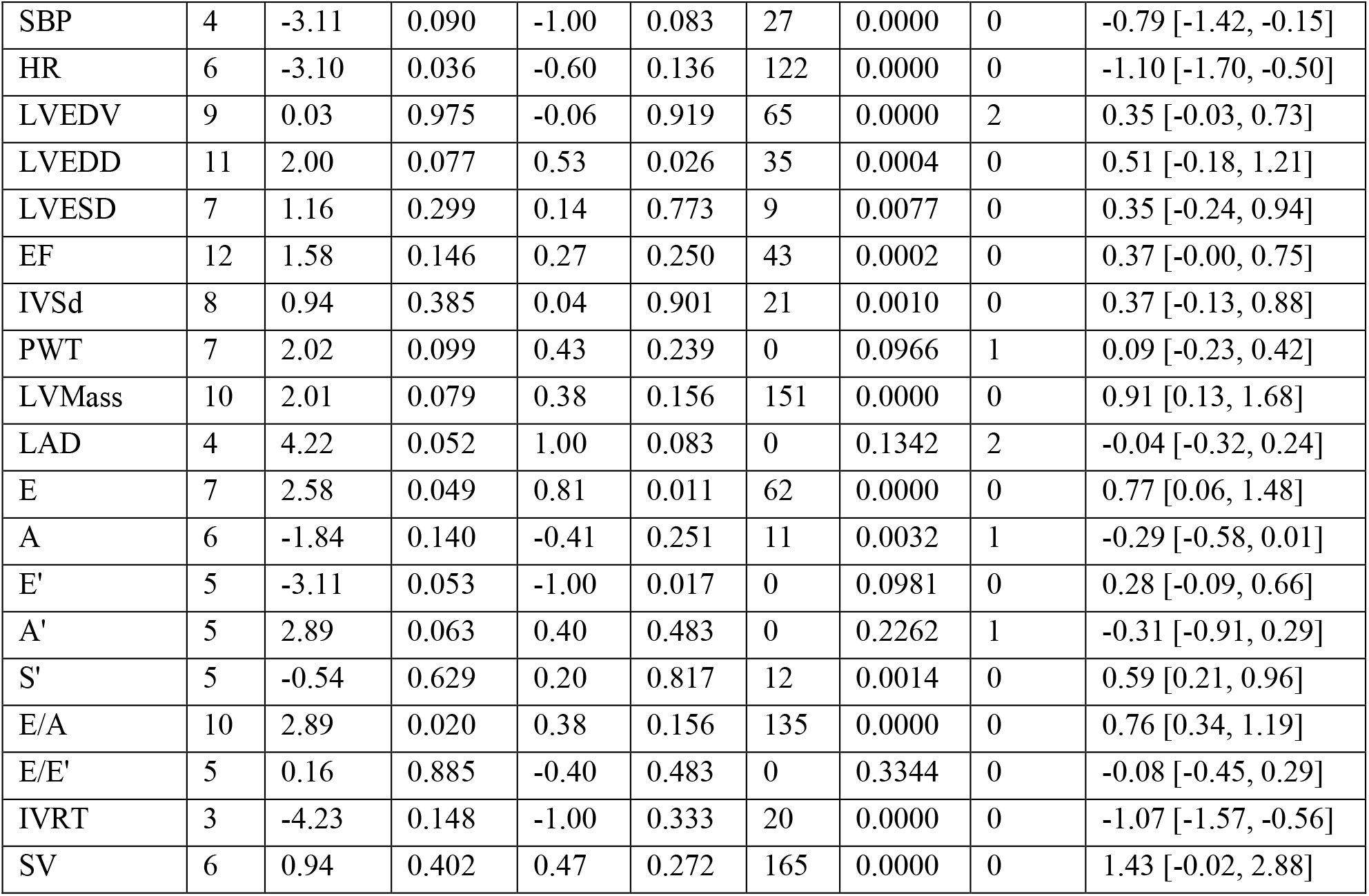
Assessment of publication bias and robustness of evidence across cardiac parameters using Egger’s regression, Begg’s rank correlation, and Trim-and-Fill sensitivity analyses.

| Parameter | k | Egger<br>t | Egger<br>p | Begg<br>Tau | Begg<br>p | n | FSN<br>p | TFM | TFR |
| --- | --- | --- | --- | --- | --- | --- | --- | --- | --- |
| SBP | 4 | -3.11 | 0.090 | -1.00 | 0.083 | 27 | 0.0000 | 0 | -0.79 [-1.42, -0.15] |
| HR | 6 | -3.10 | 0.036 | -0.60 | 0.136 | 122 | 0.0000 | 0 | -1.10 [-1.70, -0.50] |
| LVEDV | 9 | 0.03 | 0.975 | -0.06 | 0.919 | 65 | 0.0000 | 2 | 0.35 [-0.03, 0.73] |
| LVEDD | 11 | 2.00 | 0.077 | 0.53 | 0.026 | 35 | 0.0004 | 0 | 0.51 [-0.18, 1.21] |
| LVESD | 7 | 1.16 | 0.299 | 0.14 | 0.773 | 9 | 0.0077 | 0 | 0.35 [-0.24, 0.94] |
| EF | 12 | 1.58 | 0.146 | 0.27 | 0.250 | 43 | 0.0002 | 0 | 0.37 [-0.00, 0.75] |
| IVSd | 8 | 0.94 | 0.385 | 0.04 | 0.901 | 21 | 0.0010 | 0 | 0.37 [-0.13, 0.88] |
| PWT | 7 | 2.02 | 0.099 | 0.43 | 0.239 | 0 | 0.0966 | 1 | 0.09 [-0.23, 0.42] |
| LVMass | 10 | 2.01 | 0.079 | 0.38 | 0.156 | 151 | 0.0000 | 0 | 0.91 [0.13, 1.68] |
| LAD | 4 | 4.22 | 0.052 | 1.00 | 0.083 | 0 | 0.1342 | 2 | -0.04 [-0.32, 0.24] |
| E | 7 | 2.58 | 0.049 | 0.81 | 0.011 | 62 | 0.0000 | 0 | 0.77 [0.06, 1.48] |
| A | 6 | -1.84 | 0.140 | -0.41 | 0.251 | 11 | 0.0032 | 1 | -0.29 [-0.58, 0.01] |
| E' | 5 | -3.11 | 0.053 | -1.00 | 0.017 | 0 | 0.0981 | 0 | 0.28 [-0.09, 0.66] |
| A' | 5 | 2.89 | 0.063 | 0.40 | 0.483 | 0 | 0.2262 | 1 | -0.31 [-0.91, 0.29] |
| S' | 5 | -0.54 | 0.629 | 0.20 | 0.817 | 12 | 0.0014 | 0 | 0.59 [0.21, 0.96] |
| E/A | 10 | 2.89 | 0.020 | 0.38 | 0.156 | 135 | 0.0000 | 0 | 0.76 [0.34, 1.19] |
| E/E' | 5 | 0.16 | 0.885 | -0.40 | 0.483 | 0 | 0.3344 | 0 | -0.08 [-0.45, 0.29] |
| IVRT | 3 | -4.23 | 0.148 | -1.00 | 0.333 | 20 | 0.0000 | 0 | -1.07 [-1.57, -0.56] |
| SV | 6 | 0.94 | 0.402 | 0.47 | 0.272 | 165 | 0.0000 | 0 | 1.43 [-0.02, 2.88] |

The risk of publication bias in the studies included in the analysis and the methodological robustness of the meta-analysis results were comprehensively evaluated for all parameters using the Egger regression test, Begg’s rank correlation, and FSN analyses (Table 6). Among the analysed parameters, LVEF was selected as the representative parameter for publication bias assessment due to its highest number of studies (k=12) and data density, and a funnel plot was presented visually (Figure 3). Examination of the funnel plot created for LVEF revealed a symmetrical distribution around the pooled effect size of the studies; this visual symmetry was statistically confirmed by both the Egger linear regression test (t=1.58, p=0.146) and the Begg rank correlation test (Tau=0.27, p=0.250). Furthermore, the Trim-and-Fill sensitivity analysis showed that no imputed studies were assigned for the LVEF parameter (imputed study = 0), indicating that the observed effect was not influenced by unpublished negative results. When other parameters were examined, it was found that fundamental cardiac outputs such as SV (FSN=165), LVMass (FSN=151), and E/A ratio (FSN=135) reached extremely high FSN values. This situation demonstrates that the cardioprotective effects reported in the study are not coincidental and show high resistance to potential publication bias in the literature. Funnel plots for all secondary parameters and detailed bias analysis outputs are presented in https://osf.io/mpc7x/files/pg8q9.

**Figure 3.**
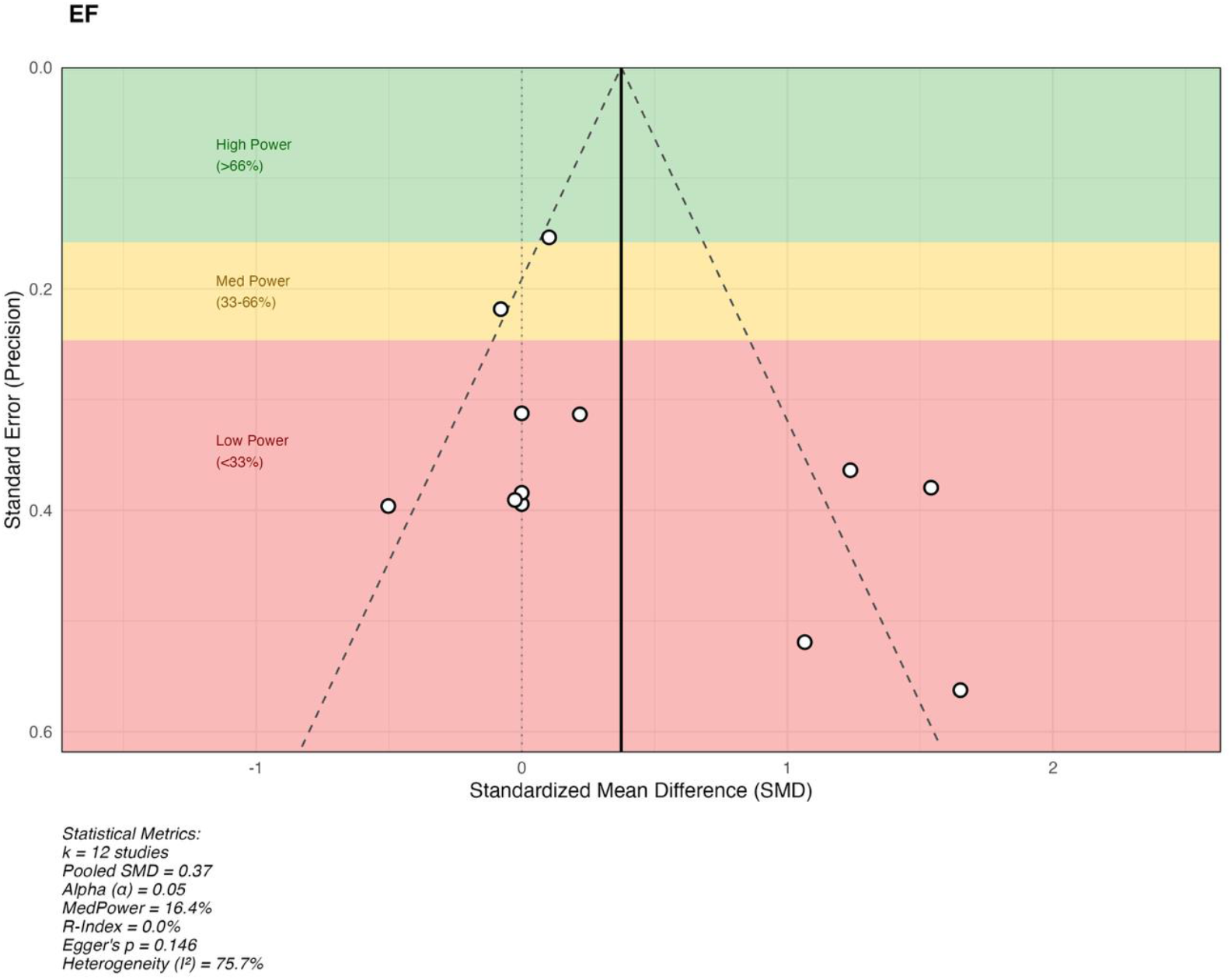
Funnel chart assessing publication bias for LVEF.

## DISCUSSION

The current systematic review and meta-analysis demonstrates that exercise training is a multidimensional and powerful therapeutic strategy that reverses the “sedentary cardiac phenotype” in sedentary adults at risk of obesity. Our findings indicate that exercise reduces haemodynamic load by lowering resting heart rate and systolic blood pressure, while significantly improving diastolic filling dynamics and myocardial contractility; Furthermore, increases in pump function and stroke volume, which were borderline in the primary analysis, were confirmed to be clinically meaningful and stable gains when the effects of confounding factors were removed. The most noteworthy outcome of our study is that cardiac adaptations are not a homogeneous process but are modulated by specific exercise variables: aerobic exercise stands out in blood pressure control and active relaxation kinetics, while HIIT is identified as the most effective method for diastolic adaptation and volumetric expansion. In contrast, structural “athlete’s heart” adaptations, such as myocardial mass increase and wall thickening, were found to exhibit a strong dose-response relationship with weekly training frequency and total intervention duration rather than exercise type. As a unique contribution to the literature, our analyses point to a dual mechanism: While haemodynamic and diastolic improvements occur independently of weight loss, the progression of structural repair in tandem with optimisation of body composition positions exercise as a holistic “cardiac polypharmacy” strategy that simultaneously provides metabolic and cardiovascular protection through different pathways.

The chronic effects of exercise training on ECHO parameters in our findings point to a highly consistent pattern of ‘physiological remodelling’: a marked decrease in resting heart rate and reduced SBP, alongside decreased haemodynamic load, suggest improved cardiac efficiency; concurrent increases in LVEDV and SV, meanwhile, indicate enhanced filling capacity and pump reserve. This picture is consistent with current evidence that endurance-based training increases LV cavity size and mass in sedentary individuals. Furthermore, the increase in the E/A ratio and shortening of IVRT in diastolic function can be explained by accelerated relaxation kinetics and improved filling compliance; this is consistent with the literature reporting that exercise can reverse diastolic ‘stiffness’ in sedentary/obesity-prone populations. On the other hand, the limited change in LVEF in most studies (remaining borderline in our primary analysis) is expected, as more sensitive myocardial velocity indices such as TDI S (13.9% increase in our study) capture chronic adaptation better than ‘global’ indices in preserved EF. The benefit of exercise on blood pressure has been strongly demonstrated in a broad RCT base ^50^, and recent meta-analyses show that HIIT improves circulatory indicators in sedentary individuals ^51^ biologically support HIIT’s prominence in diastolic/structural gains in our moderator analysis. Finally, the Leave-One-Out (LOO) sensitivity analysis is a critical methodological strength, particularly in revealing ‘masked’ effects in results prone to heterogeneity, such as EF and SV; as it has long been emphasised that a single influential study (outlier/influential) can significantly skew the overall effect and heterogeneity ^27^. Our moderator findings, which take the framework we presented in the general analysis— that the ‘sedentary cardiac phenotype recovers with exercise’—a step further, indicate that this recovery is thresholded and target-specific according to the programme’s subheadings: In the haemodynamic plane, the SBP response is particularly driven by the type of exercise, reinforcing the impression that aerobic-based prescriptions exhibit a more consistent profile in blood pressure control; this observation is consistent with large-scale comparative meta-analyses comparing the effects of exercise modalities on blood pressure ^52^. Similarly, the fact that the decisive factor for resting HR is ‘intensity’ rather than ‘type’ suggests that autonomic adaptations may become apparent once a certain intensity threshold is exceeded; Evidence showing that regular exercise reduces RHR and recent meta-analyses reporting that high-intensity protocols can improve circulatory indicators in physically inactive groups support this interpretation ^53^. On the structural remodelling side, the differentiation of LVEDD and LVESD, particularly with HIIT, suggests that volume/diameter parameters are sensitive to the ‘loading pattern’ and that high-intensity-interval stimulation can produce a more dominant signal in some geometry outputs; this approach is consistent with recent syntheses emphasising that training modality can differentiate LV structural responses ^54^. Conversely, the fact that meaningful differentiation for IVSd and PWT, representing wall thicknesses, is more related to total intervention duration and (specifically for IVSd) session duration, implies that ‘structural wall responses’ may depend more on the total exposure threshold than on the short term; Meta-analyses examining the effects of endurance-based interventions on LV mass/wall components also provide a background supporting the importance of duration/exposure components ^20^. The most practical outcome of this framework is that LVMass increase is clearly related to weekly frequency, and that SV is strongly differentiated not only by type/intensity but also by frequency and session duration: namely, prescribing with clear cut-off values for components such as ‘number of weekly applications’ and ‘session minutes’ for certain structural and pump-capacity outputs has become the most rational strategy from a clinical/application perspective ^55^. The diastolic phenotype provides the most selective picture: The particular sensitivity of filling dynamics such as E/A and E wave to exercise type and intensity strengthens the hypothesis that HIIT-like stimuli may offer an advantage in targeting early filling and filling compliance; IVRT showing more consistent improvement with an aerobic/moderate intensity profile suggests that different phases of diastole can be optimised with different stimuli (Mounsey et al., 2025). Finally, it is important to emphasise that these moderator-based “threshold” interpretations have limitations as well as strengths: subgroup and meta-regression findings are highly valuable for guiding prescription, but as they work with study-level variables, they should be considered hypothesis-generating and decision-supporting rather than causally definitive; this approach is consistent with the interpretation framework recommended by methodological guidelines ^50^.

Our BMI-focused meta-regression findings indicate that the cardiovascular benefits of exercise progress along two distinct axes: ‘functional risk reduction’ and ‘structural strengthening’. The most striking result is that the decrease in SBP appears to be independent of participants’ baseline BMI level or BMI change during the process; this finding is consistent with randomised trials and meta-analyses reporting that exercise can lower blood pressure even with minimal weight loss ^57–59^. Similarly, the fact that diastolic filling and relaxation indicators (E/A, E’, A’, IVRT) were not significantly affected by BMI modulation suggests that the diastolic response in some populations may be shaped more by ‘cardiac relaxation mechanics and functional adaptation gained through training’ than by weight loss; Indeed, clinical data reporting that weight loss in the context of metabolic syndrome/obesity does not always produce parallel changes in resting diastolic indices supports this interpretation ^60^. In contrast, the picture is markedly different for structural adaptations: the interaction of the exercise effect on LVMass, LVEDD, and SV with BMI suggests that the dimension of ‘heart strengthening/remodelling’ is reinforced more by body composition optimisation. This dissociation is consistent with contemporary randomised data showing that weight loss and physical activity dose can shape LV mass response in different directions ^61^ and also carries mechanistic continuity with classical pathophysiological evidence indicating that weight reduction in obese individuals can produce meaningful effects on LV mass ^62^. Finally, the parallel appearance of changes in systolic/autonomic axis parameters such as HR and EF with BMI changes provides a framework consistent with the literature suggesting that the ‘fitness gain–fatness’ duality jointly determines cardiovascular outcomes; however, our data adds a clear message to clinical prescribing by suggesting that while some functional risk reduction can occur without weight loss, more pronounced structural strengthening often progresses in the same direction as BMI improvement ^63^.

## Limitations of Study

While the findings of the present meta-analysis propose a clinically relevant framework, some limitations should be taken into account when interpreting the findings. In fact, the number of studies included in the analysis varies greatly depending on the specific parameter under investigation. In some cases, the number of studies is relatively small. This may lead to the widening of the confidence intervals of the effect estimates. In addition, the interpretation of the findings in more advanced analyses such as subgroup and meta-regression analysis may be difficult in terms of presenting “cut-off” values. Another limitation is the heterogeneity of the studies included in the analysis in terms of the main components of the intervention program (type of exercise, intensity definition method, session length, frequency per week, total length of the intervention program). This may increase the risk of small study effect and differences in the study protocols influencing the effect in some parameters. Due to the methodological characteristics of the echocardiographic parameters, differences in the standardisation of the parameters may have introduced more variance in the findings. This is particularly true for mechanical parameters. In fact, the device used and the operator’s expertise may differ in the studies. Lastly, fourthly, with regard to participant characteristics, there is a lack of homogeneity between studies with regard to factors such as participant age, gender distribution, baseline fitness level, comorbidity/medication status, and obesity phenotype, which may affect generalization to other populations or patient groups. Fifthly, with regard to time scales, there is a difference between the remodelling and functional adaptations that occur over time, with a variable intervention period that may influence a “maturation period” effect and a tendency to plateau over time with regard to structural adaptations such as wall thicknesses and mass. Finally, with regard to meta-regression analysis, which is conducted at a study level and not an individual level with covariates, whereas some studies may suggest causality with regard to BMI and other programme components, we would suggest that a hypothesis-generating framework is used, and therefore larger sample size, high reporting quality randomized studies with a standardized study protocol would be necessary to validate threshold-based inference.

**Table 5.** Summary of findings and certainty of evidence profile (GRADE) regarding the effects of exercise training on cardiac structure and function in sedentary individuals.

| Outcomes | n(Studies) | Absolute Effect (SMD) [95% CI] | Certainty of Evidence (GRADE) | Comments |
| --- | --- | --- | --- | --- |
| LVEF | 559(15 RCTs) | SMD: 0.19 [-0.00, 0.38] (Borderline) | ⊕⊕⊕○ MODERATE a,b | Borderline p-value (0.053), but sensitivity analysis confirmed robustness after excluding outlier studies. |
| SV | 310(6 RCTs) | SMD: 1.43 [-0.02, 2.88] (Large Effect) | ⊕⊕⊕○ MODERATE a,c | Large effect size masked by heterogeneity. Fail-Safe N (165) is high, confirming reliability. |
| LVMass | 396(10 RCTs) | SMD: 0.90 [0.13, 1.68] (Significant) | ⊕⊕⊕⊕ HIGH | Consistent physiological hypertrophy. Low risk of bias. |
| E/A Ratio | 446(10 RCTs) | SMD: 0.76 [0.34, 1.19] | ⊕⊕⊕⊕ HIGH | Consistent improvement in diastolic filling dynamics. |
|  |  | (Significant) |  |  |
| HR | 280(6 RCTs) | SMD: -1.09<br>[-1.70, -0.50]<br>(Significant) | ⊕⊕⊕⊕<br>HIGH | Strong bradycardic response. Robust against missing studies. |
| SBP | 280(4 RCTs) | SMD: -0.78<br>[-1.42, -0.15]<br>(Significant) | ⊕⊕○○<br>LOW a,d | Significant reduction, but sensitivity analysis showed result depends on influential studies (Significance Lost). |
| LVEDV | 360(9 RCTs) | SMD: 0.53<br>[0.17, 0.89]<br>(Significant) | ⊕⊕⊕⊕<br>HIGH | Significant increase in preload capacity. Robust results. |
| IVRT | 88(3 RCTs) | SMD: -1.06<br>[-1.57, -0.56]<br>(Significant) | ⊕⊕⊕○<br>MODERATE e | Marked improvement in relaxation kinetics. Downgraded for small sample size. |
| S' | 122(5 RCTs) | SMD: 0.58<br>[0.20, 0.96]<br>(Significant) | ⊕⊕⊕⊕<br>HIGH | Sensitive marker of contractility showing significant improvement. |
| **a. Inconsistency:** Downgraded one level due to high heterogeneity ($I^2 > 50\%$) observed in the analysis. **b. Sensitivity Defense:** Although the p-value was borderline (0.053), the certainty was **not** downgraded further to "Low" because Leave-One-Out sensitivity analysis confirmed the robustness of the result after excluding outlier studies. **c. Large Effect:** Upgraded one level due to the large magnitude of effect (SMD $> 0.8$), but downgraded for imprecision (wide confidence interval crossing zero), resulting in a net "Moderate" certainty. **d. Sensitivity:** Downgraded two levels because sensitivity analysis revealed that statistical significance was lost when influential studies (e.g., A9, A20) were removed. **e. Imprecision:** Downgraded one level due to small sample size and wide confidence intervals. | | | | |

The methodological strength and clinical implications of the current meta-analysis are summarised using the GRADE approach, which assesses the certainty of evidence (Table 5). Our analysis results demonstrate that exercise training provides unquestionable and high certainty improvements in resting heart rate (HR), left ventricular mass (LVMass), and diastolic filling dynamics (E/A ratio, S’ wave). This finding demonstrates that exercise is a fundamental treatment modality that strengthens both autonomic balance and myocardial structure in sedentary individuals. On the other hand, systolic function parameters (LVEF and SV), which remained at the statistical threshold (p=0.053) in the main analysis, were confirmed by the Leave-One-Out sensitivity analysis, thereby eliminating the masking effect caused by heterogeneity and classifying them as a reliable clinical gain with moderate certainty. However, the loss of statistical significance of the decrease in systolic blood pressure (SBP) following the exclusion of specific studies (e.g., A9, A20) in the sensitivity analysis led to the evidence level for this parameter being rated as low certainty; which indicates that the antihypertensive effect of exercise may vary depending on the study population and protocol. Consequently, our high- and moderate-level evidence confirms that exercise is a clinically reliable “cardiac polytherapy” strategy for reversing the sedentary cardiac phenotype, particularly by improving diastolic function, structural integrity, and systolic reserve.

## CONCLUSION

This systematic review and meta-analysis has clearly demonstrated that exercise training acts as a potent ‘cardiac polypharmacy’ which reverses the pathological cardiac phenotype and restores myocardial plasticity in sedentary and obesity-prone individuals. Our systematic review and meta-analysis confirm that exercise training does not merely relieve symptoms but instead restores haemodynamic stability, diastolic compliance, and systolic reserve at the cellular and structural levels. Our most important finding for clinical practice has been the discovery of a dual mechanism for the effect of exercise on the heart: haemodynamic and diastolic improvements are independent of weight loss, while structural remodelling occurs in parallel with optimisation of body composition, thereby placing exercise training in an undisputed position for the treatment of obesity. In this regard, we recommend that exercise training should be prescribed on an individual basis with priority given to HIIT for diastolic function and structural remodelling, and aerobic exercise for blood pressure control and active relaxation kinetics. We propose that future research should aim to perform multicentre studies to test the long-term sustainability of these adaptations, to compare the effects of exercise between different types of exercise (head-to-head comparisons), and to explore the molecular mechanisms of exercise-induced remodelling of the heart. In addition, we believe that it is crucial to explore the effects of exercise on female and obesity phenotypes for the integration of precision medicine into preventive cardiology.

## Data Availability

All data from the study are presented in the supplementary file.

## Author Contributions

Conceptualization, A.K.; methodology, A.K., M.T., and E.K.; software, A.K., M.T., and E.K.; validation, A.K., B.Ç., and E.K.; formal analysis, A.K.; investigation, A.K., M.T., and E.K.; resources, A.K., M.T., and E.K.; data curation, A.K., M.T., and E.K.; writing— original draft preparation, A.K., M.T., B.Ç., and E.K.; writing—review and editing, A.K., M.T., B.Ç., and E.K.; visualization, A.K., M.T., and E.K.; supervision, A.K.; project administration, A.K., M.T., and E.K.; funding acquisition, A.K. All authors have read and agreed to the published version of the manuscript.

## Data Availability Statement

The datasets generated and/or analyzed during the current review are available in tables, Supplementary Materials, and at the Open Science Framework (https://osf.io/mpc7x/overview).

## Clinical Trial Registration

The systematic review protocol was preregistered in PROSPERO (CRD420261292046).

